# Understanding adherence to self-isolation in the first phase of COVID-19 response

**DOI:** 10.1101/2022.03.14.22272273

**Authors:** Charlotte Robin, Rosy Reynolds, Helen Lambert, Matthew Hickman, G James Rubin, Louise E Smith, Lucy Yardley, Shenghan Cai, Tingting Zhang, Piers Mook, Oliver McManus, Gemma Lasseter, Polly Compston, Sarah Denford, Juan Zhang, Richard Amlôt, Isabel Oliver

**Author notes:** Corresponding author: Charlotte Robin.

## Abstract

**Objective:** To gain a better understanding of decisions around adherence to self-isolation advice during the first phase of the COVID-19 response in England.

**Design:** A mixed-methods cross sectional study.

**Setting: England:** Participants COVID-19 cases and contacts who were contacted by Public Health England (PHE) during the first phase of the response in England (January-March 2020).

**Results:** Of 250 respondents who were advised to self-isolate, 63% reported not leaving home at all during their isolation period, 20% reported leaving only for lower risk activities (dog walking or exercise) and 16% reported leaving for potentially higher risk, reasons (shopping, medical appointments, childcare, meeting family or friends). Factors associated with adherence to never going out included: the belief that following isolation advice would save lives, experiencing COVID-19 symptoms, being advised to stay in their room (rather than just “inside”), having help from outside and having regular contact by text message from PHE. Factors associated with non-adherence included being angry about the advice to isolate, being unable to get groceries delivered and concerns about losing touch with friends and family. Interviews highlighted that a sense of duty motivated people to adhere to isolation guidance and where people did leave their homes, these decisions were based on rational calculations of the risk of transmission – people would only leave their homes when they thought they were unlikely to come into contact with others.

**Conclusions:** Measures of adherence should be nuanced to allow for the adaptations people make to their behaviour during isolation. Understanding adherence to isolation and associated reasoning during the early stages of the pandemic is an essential part of pandemic preparedness for future emerging infectious diseases.

**Strengths and limitations of this study:**

- Our participants were contacted directly by Public Health England during the first three months of the pandemic – the only cohort of cases and contacts who experienced self-isolation during this early phase of the pandemic.
- Results may not be directly generalisable to wider populations or later phases of pandemic response.
- We classified reasons for leaving the home as higher or lower contact, as a proxy for potential risk of transmission, however further research published since we conducted our research as refined our understanding of transmission risk, highlighting the need for more in-depth research on adherence behaviour and transmission risk.
- The mixed methods approach combined quantitative measures of adherence with an exploration of how and why these decisions were being made in the same people.
- Our study provides unique insights into self-isolation during the earliest stages of the pandemic, against a background of uncertainty and lack of information that will recur, inevitably, in the face of future pandemic and similar threats.

## Introduction

In response to the global COVID-19 pandemic, governments around the world have placed great importance on test, trace and isolate systems as a strategy to minimise transmission^1^. In the United Kingdom (UK) the isolation of people with symptoms and their contacts was vital in the early ‘containment’ phase of the pandemic response (January to March 2020), particularly before widespread testing was available. As a public health measure, isolation aims to prevent person-to-person spread of infections by separating people to interrupt transmission^2^. For COVID-19, this includes separating exposed from unexposed individuals, because of evidence of asymptomatic transmission^3,4^. Adherence to these measures is essential to limit community transmission of the virus.

A growing body of evidence indicates variation in the extent to which people adhere to self-isolation guidance and what factors may influence adherence^5,6,7,8^. However, understanding adherence to self-isolation is limited by how adherence is measured. There are no validated measures of adherence to self-isolation and generally adherence is measured as a self-reported binary outcome; adherent or not^5^. While measuring adherence in a binary way is useful for determining changes in adherence over time and providing rapid and pragmatic insights into behaviour, how individuals understand and adhere to self-isolation is likely to be more nuanced^7,9,10^. It is also unclear how the public negotiate decisions around adherence to self-isolation guidance in the context of contact tracing, specifically when that advice has been provided directly to individuals by public health agencies.

During the first phase of England’s COVID-19 response (January to March 2020), contact tracing was managed by Public Health England (PHE), prior to the launch of the national NHS Test and Trace service in May 2020. Regional Health Protection Teams at PHE aimed to contact all known cases and their contacts to advise them of their status, provide them with information on self-isolation guidance and offer them support during their isolation period. Adherence to self-isolation during this phase and how people were making those decisions has not previously been determined.

The aim of this study was to gain a better understanding of adherence to self-isolation advice in cases and contacts who were identified through contact tracing in England during the first phase of the pandemic response, when anxiety levels in the general population were higher than normal^11,12^. Understanding factors affecting adherence during these initial phases of the pandemic response is particularly important as high adherence to isolation will give the best chance of containing the virus before community transmission becomes widespread, and future emerging infectious disease outbreaks will be characterised by similar high uncertainty and high caution. Regardless of whether future novel pathogens are harmless, the initial stage of any pandemic response is likely to focus on containment.

## Methods

In early 2020, details of all cases and contacts who were contacted by PHE’s Health Protection Teams were recorded on PHE’s case management system (HPZone). All participants were sampled from this system and invited to take part in an online survey and follow-up qualitative interview.

### Case definitions for survey inclusion

Confirmed cases had a PCR positive test for SARS-CoV-2. Possible cases had a history of exposure (to a confirmed case or by reason of travel history) and symptoms of fever or dry cough or breathing difficulty. Contacts were people exposed to a confirmed case. For the purposes of our survey, we classified an individual based on the circumstances which would have prompted first contact from PHE.

### Sampling

All cases and contacts (as defined above) in England aged 18 years or over and entered onto PHE’s case management system ‘HPZone’ by 12^th^ March 2020 were potentially eligible. After applying exclusion criteria (Table S1 – supplementary information), 350 confirmed cases, 1472 possible cases and 1794 contacts were invited to participate in the survey.

### Survey

An online survey (supplementary information) was developed using Snap Survey v11 (Snap Surveys, Bristol, UK), including sections on sociodemographic and household characteristics, self-reported adherence to advice received, self-reported barriers and facilitators to following advice, and a self-assessment of mental health and wellbeing using standardised tools. Mental health outcomes will be reported separately. The survey was piloted among 15 cases and 15 contacts, and minor changes to wording were made to improve clarity.

### Interviews

Semi-structured interviews were used to explore experiences of self-isolation in more depth. A topic guide with open-ended questions was used to ensure key areas were covered but was used flexibly to allow exploration of new themes as they arose. The topic guide included sections on experiences of self-isolation, adherence to guidance, seeking information, advice and support. Interviews took place over the telephone or Skype, were audio recorded and transcribed *verbatim*.

### Recruitment

#### Survey

The survey was completed in two phases with invitations sent on 24^th^ July 2020 and 9^th^ October 2020. The first phase invited 463 cases (232 confirmed, 231 possible) and 451 contacts; the second phase invited all remaining eligible cases (118 confirmed, 1241 possible) and contacts (1343).

Invitations were sent via SMS, including a link to an online participant information sheet and the survey. A follow-up reminder SMS message was sent after 3-4 weeks; if no response was received after a further week, the invitee was recorded as a non-responder and no further contact was made. The survey could be completed anonymously, but respondents who consented to participate in voluntary follow-up qualitative interviews were asked to provide their contact details.

#### Interviews

Respondents who consented to interview were randomly selected to take part, stratified by status (case or contact) and index of multiple deprivation (IMD) quintile, based on home postcode. Overall, 78 respondents consented to interview, of whom 30 were invited and 16 interviews took place, between July and November 2020.

### Analysis

#### Survey

Analysis used Stata v15.1 (Stata Statistical Software 2017; StataCorp LLC, College Station, TX). Categorical data were described by percentage. Age was described by median and interquartile range; other continuous data were described by mean or categorised.

The primary measure of adherence was staying at home for the full duration when advised to do so. Respondents reported how often they left home for various reasons (Table 1) and responses were dichotomised as ever *versus* never for analysis. We categorised reports of leaving home during the isolation period into lower and higher-contact outings, defining exercise and dog-walking as lower-contact and all other reasons (listed in Table 1) as higher-contact. The survey did not distinguish between outdoor and indoor exercise, but respondents who left home only to exercise or walk dogs reported little indoor contact with others away from their home suggesting that exercise was largely outdoors.

**Table 1.** Adherence to staying at home.

| Reason<sup>2</sup> | Left home for given reason and frequency: N (%)<sup>1</sup> |  |  |  |  |
| --- | --- | --- | --- | --- | --- |
|  | Missing | Not applicable or not at all | Occasionally | More than half the days | Nearly every day |
| Shop – essential | 0 | 225 (90.0) | 23 (9.2) | 2 (0.8) | 0 |
| Shop – other | 1 (0.4) | 248 (99.2) | 1 (0.4) | 0 | 0 |
| Exercise | 0 | 187 (74.8) | 27 (10.8) | 10 (4.0) | 26 (10.4) |
| Medical | 0 | 233 (93.2) | 17 (6.8) | 0 | 0 |
| Work | 0 | 250 (100) | 0 | 0 | 0 |
| Childcare/school | 0 | 247 (98.8) | 3 (1.2) | 0 | 0 |
| Help someone else | 0 | 250 (100) | 0 | 0 | 0 |
| Meet people | 0 | 246 (98.4) | 3 (1.2) | 1 (0.4) | 0 |
| Walk dog | 0 | 230 (92.0) | 7 (2.8) | 4 (1.6) | 9 (3.6) |

<sup>1</sup> Excludes 72 respondents who did not report being advised to isolate and so were not asked about going out.
<sup>2</sup> See copy of survey (Q26) in supplementary information for full wording
Two responses citing 'Another reason' have been recoded into the categories above: "To have a covid test" is shown as 'Medical' and "Only to take the rubbish out and collect items left on doorstep" is treated as not going out at all.

We explored potential associations of 62 factors from the survey (Table S4) with adherence in two ways. Firstly, we described the overall pattern of leaving home using three categories – never going out, going out only for lower-contact reasons, going out for higher-contact reasons – and tested for association with categorical factors by Fisher’s exact test; for multi-level factors, we repeated this with dichotomised forms for comparison with the second approach. Secondly, we considered three behaviours as separate binary outcomes – never (vs ever) going out for any reason, ever (vs never) going out for lower-contact reasons, ever (vs never) going out for higher-contact reasons – and, for each behaviour, estimated the risk ratio (RR) and p-value for each (dichotomised) factor. To identify differential effects of factors on lower- and higher-contact outings, we used seemingly unrelated estimation to compare the estimated risk ratios.

We took p≤0.1 as ‘some evidence’ for association or differential association.

#### Interviews

Transcripts were coded using an open approach i.e. codes were not decided *a priori*. This process disassembled data into discrete parts to develop a list of codes. Memos on emerging ideas and possible relationships between codes were kept alongside initial codes and codes that represented similar concepts were assembled into conceptual categories. Coding was performed iteratively within and between transcripts, using the technique of constant comparative analysis. The constant comparison between data and analysis allowed the development of codes, categories and theories to be tested across transcripts^13^ until a final coding framework was developed. This coding framework was also applied to the 200 free-text comments from the survey; no additional codes were developed during this phase of the analysis.

## Findings

The overall response rate for the survey was 9% (322/3616), including 52 confirmed cases, 91 possible cases and 179 contacts. Survey participant and invitee characteristics are shown in Table S2. Characteristics of interview participants are shown in Table S3.

Overall, of the 250 survey respondents who had been advised to self-isolate most reported adhering to the advice (Table 1); 158 (63%) reported not leaving home at all during their isolation period and 51 (20%) reported leaving only for lower-contact activities i.e. exercise or dog walking. Sixteen percent (41) left home for higher-contact reasons: shopping, medical appointments, childcares or meeting family and friends. Five (2%) had occasional visitors to their homes.

Reasons for leaving the home during isolation were grouped into two categories based on contact with other people – lower contact and higher contact. Evidence from the survey supported the classification of exercise and dog-walking as lower-contact, implying lower-risk, activities than leaving home for any other reasons (listed in Table 1). The 51 people who left home only for dog-walking or exercise reported less contact with other people away from their own homes, compared with the 41 who went out for other reasons: only 18% vs 46%, respectively, ever spent time with people indoors, keeping >2 m away; 10 vs 27% had closer indoor contacts; and 14 vs 51% had to touch surfaces other people had touched. We found evidence that some factors had different patterns of association with lower- and higher-contact outings, indicating that respondents distinguished between them (Table 2).

**Table 2.** Factors showing some evidence (p≤0.1) of association with going-out behaviours.

| Q | Factor<sup>1</sup> | Behaviour<sup>2</sup> |  |  | 3-level outcome<sup>3</sup> (p) |
| --- | --- | --- | --- | --- | --- |
|  |  | Never out | Lower-contact outings | Higher-contact outings |  |
| Little or no evidence of difference in RRs for lower- and higher-contact outings |  |  |  |  |  |
| Q44d | Agree: following advice would save lives | ↑ | ↓ | ↓ | (0.03) |
| HPZ | Survey phase 2 | ↑ | ↓ | ↓ | (0.07) |
| Q53 | Angry about being asked to self-isolate | ↓ | ↑ | ↑ | (0.02) |
| Q44m | Had help from outside | ↑ | ↓ | ↓ | (0.04) |
| Q12 | Fever, dry cough or breathing difficulty<sup>4</sup> | ↑ | ↓ | ↓ | (0.09) |
| Q56 | Ethnic group (all non-White ethnic groups) | ↑ | ↓ | ↓ | (0.04) |
| Q21 | Home had a room I could live/sleep in | ↓ | ↑ | ↓ | (0.07) |
| Some evidence (p≤0.1) of differing RRs, association appears stronger for lower-contact outings |  |  |  |  |  |
| Q7 | Advised to stay in room (vs stay inside) | ↑ | ↓ | ↓ | (0.06) |
| Q54 | Age ≥50 | ↓ | ↑ | ↓ | (0.04) |
| Q48c | Had help with pets (if had any pet) | ↑ | ↓ | ↑ | (0.05) |
| Q22 | Home had some outside space | ↓ | ↑ | ↓ | (≤0.01) |
| Some evidence (p≤0.1) of differing RRs, association appears stronger for higher-contact outings |  |  |  |  |  |
| Q30 | Tried but unable to get groceries delivered | ↓ | ↑ | ↑ | (≤0.01) |
| Q44o | Mental health worsened | ↓ | ↓ | ↑ | (0.01) |
| Q44a | Would lose touch with family/friends | ↓ | ↑ | ↑ | (0.01) |
| Q32d | Very ill, needed care from family | ↓ | ↓ | ↑ | (0.01) |
| Q44n | Physical health worsened | ↓ | ↓ | ↑ | (0.02) |
| Q23 | Had a pet at home | ↑ | ↓ | ↓ | (0.06) |
| Q9a | PHE contact by text (vs email or phone) | ↓ | ↑ | ↓ | (0.07) |
| Q44i | I could pass virus on if I went out | ↓ | ↑ | ↓ | (0.05) |

<sup>1</sup> The full wording of the relevant questions is in the survey, in supplementary material.
<sup>2</sup> The three 'behaviour' analyses treated each behaviour individually as a binary outcome: never (vs ever) going out; any (vs no) lower-contact outings; any (vs no) higher-contact outings.
<sup>3</sup> The 'three-level outcome', the primary analysis, considered the pattern of respondents' going-out behaviour as a whole, classified as: never / lower-contact only / higher-contact (with or without lower-contact as well).
<sup>4</sup> Symptoms recognised at the time as indicating Covid-19.
↑ Some evidence of association ( $P \leq 0.1$ ): a higher % of those with the factor report the behaviour ( $RR > 1$ ).
↑ Little/no evidence association ( $P > 0.1$ ) but point estimate of $RR > 1$ .
↓ Some evidence of association ( $P \leq 0.1$ ): a lower % of those with the factor report the behaviour ( $RR < 1$ ).
↓ Little/no evidence of association ( $P > 0.1$ ) but point estimate of $RR < 1$ .
Arrow colours: green=greater adherence; purple=lower adherence.
RR=risk ratio.

We found some evidence of association with the overall pattern of adherence – never going out, going out only for lower-contact reasons, going out for higher-contact reasons – for 19 dichotomised factors (Table 2). Those that relate to observations from the qualitative interviews are described in more detail below, alongside those insights.

### Shift in identity

During the early phase, all contact tracing was conducted by Health Protection Teams and therefore all participants were contacted directly by Public Health England (PHE) to inform them of their status as either a case or contact. Of 322 survey respondents, 43 (13%) recalled the reason for PHE contact being to inform them of a positive test result, 72 (22%) to ask about symptoms and arrange testing and 152 (47%) to inform of contact with a case (25 within their household and 127 outside). Of those who reported receiving advice, 204/250 were advised to “stay inside” and 46 to “stay in my room”.

The interviews revealed that receiving this contact resulted in participants experiencing a sudden shift in their identity, unexpectedly being classified as a case or a contact. This new identity brought with it certain rules and restrictions that they had to abide by. Their social world had abruptly become very different (Table 3a, Quote 1).

**Table 3a.**
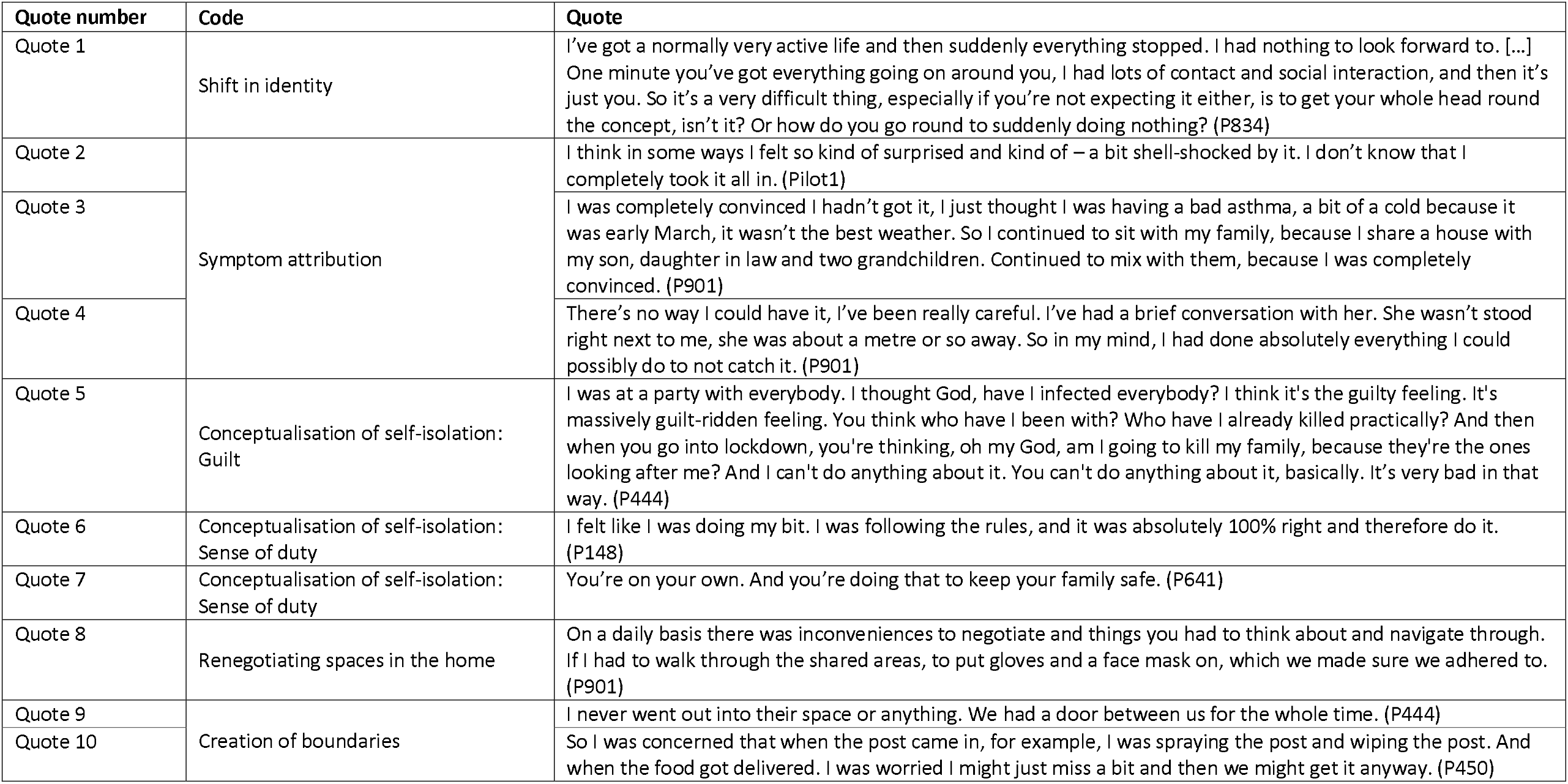
Experiences of contact tracing and self-isolation.

**Table 3b.**
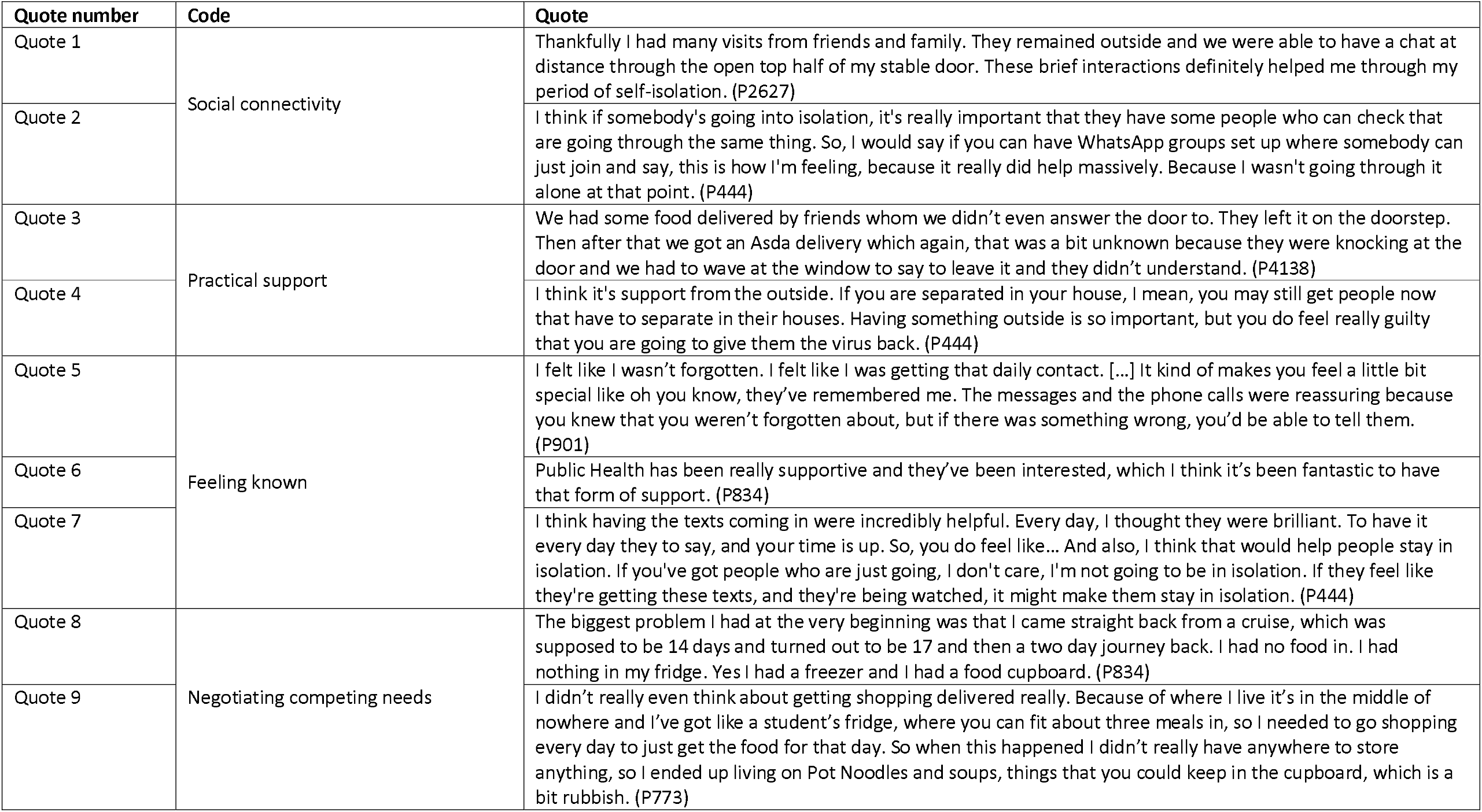
Adherence to self-isolation.

**Table 3c.** Rational adaptations to mitigate risk.

| Quote number | Code | Quote |
| --- | --- | --- |
| Quote 1 | Rational adaptations to mitigate risk | Daily early morning walk or evening when no one is around helps to stay positive. Obviously this would depend on where you live but [town], as it is quite spread out, makes it easier to exercise outside while still staying away from others. (P1278) |
| Quote 2 |  | I think the only time that we left the house, other to go in our own garden. We went out once in the car. Just to escape the four walls. We stayed inside the car and just drove round the countryside for a short while and came back again. (P374) |
| Quote 3 |  | Well, it's the fact that he [the dog] wants to go out. To walk. If we didn't stick totally to the go out for exercise once a day, then I would've found that difficult with him. I was going out 6:00 o'clock in the morning. Taking a good walk. Not seen a soul. And then taking him out around a bit later on and avoiding anybody you saw. If you saw anybody, it was the odd person. That was it. So, I must admit, I broke the rule with that. (P621) |
| Quote 4 |  | I did, during my isolation, if I'm really honest, because we've got a dog, I would take him. I drove somewhere where I knew that I wouldn't see other people and took my dog for a walk. And did that, you know, because it was only fair to do that. But it was really trying to keep away from other people. (P327) |
| Quote 5 | Over-adherence | It's made me less inclined to go out. I'm definitely less inclined to go out. I think you develop a bit of a safety bubble, whether it's consciously or not. And you just know that your home is your bubble. So it makes you less likely to want to expose yourself. It definitely makes you sub-consciously create your own space and not necessarily want anything to penetrate that. You want to stay very much where you know you're safe. (P901) |
| Quote 6 |  | I suppose a bit nervous. Not nervous, that's not the right word, that kind of like apprehensive feeling. It doesn't take long to create habits, you know, like I suppose two weeks had felt like long enough for it to feel a bit overwhelming when we went outside. (P808) |
| Quote 7 |  | I think in some ways it has made is kind of sealed off and reluctant to get back to some sort of normal. So we're tending to keep our little sealed bubble going for now. (P901). |

### Symptom attribution

For cases, the unexpected nature of the shift in their identity was related to how they conceptualised their symptoms. In some instances, despite knowing the case definition and having known exposure to a case or recent travel to a high-incidence country, there was still a sense of disbelief that the symptoms they were experiencing were actually COVID-19 (Table 3a, Quote 2 and 3). One of these participants was also aware that a colleague had recently returned to work after visiting a high-incidence country, yet felt that the precautions she took in the workplace meant the symptoms she subsequently experienced could not be COVID-19 (Table 3a, Quote 4).

This sense of uncertainty eased once cases were able to access testing. Of the 322 survey respondents, 96 reported definitely having had COVID-19, with 82 having this confirmed by a test. There was a high degree of trust in a remembered positive test result, 82/88 believing that they had definitely had coronavirus. However, there was stronger evidence of association with behaviour for symptoms than for a positive test, or report of having had COVID-19. The 95 respondents who remembered having fever, dry cough or breathing difficulty were less likely than others to have left home for lower-contact reasons (20% vs 33%), but there was no evidence of a difference in higher-contact outings.

### Conceptualisation of self-isolation

A part of the sudden shift in identity in becoming a case or contact was the realisation that they were now potentially a vector for the virus. In some cases, participants felt a sense of guilt over the potential risk of transmission and harm to others (Table 3a, Quote 5).

#### Sense of duty

In addition to feeling guilty, participants accepted self-isolation as a way of mitigating risk of further transmission. They felt a sense of duty to protect others and this helped them to adhere to guidance (Table 3a, Quote 6 and 7). This sense of duty was reflected in the survey, where respondents advised to self-isolate (212/250, 85%) agreed or strongly agreed that following the advice would help save lives. This belief was associated with greater adherence to self-isolation guidance and fewer reported outings for any reason (33 vs 55%). Similarly, the majority of survey respondents (87%) agreed or strongly agreed that following advice to self-isolate would help to protect the NHS; however there was no evidence that this belief was associated with adherence to the advice received.

#### Renegotiating spaces in the home

One way in which participants managed the risk they posed to people they lived with was through renegotiating spaces within their home. Following being identified as a case or contact, spaces within their home were subsequently designated clean or contaminated, which now had to be taken into consideration in their day-to-day lives, for example using personal protective equipment in shared spaces (Table 3a, Quote 8). Similar behaviours were reflected in the survey respondents. Of 250 respondents advised to isolate, 76% reported washing their hands “nearly every time”, whereas only 48% of respondents reported cleaning surfaces and objects at the same frequency. Among people advised to stay inside (not in their room), such frequent handwashing was reported more often by people who lived with others (125/161, 78%), compared with those who lived alone (27/43, 63%).

#### Creation of boundaries

To help negotiate these contaminated spaces in their homes, participants created boundaries to reduce the risk of transmission, for example ensuring a barrier between the designated clean and contaminated spaces (Table 3a, Quote 9). As well as keeping the virus within the confines of the home (or within certain spaces within the home), for some participants the boundary around the home had the dual purpose of keeping the virus out. For them, everything outside the home was potentially contaminated and they enacted a strict hygiene routine to try and minimise contamination (Table 3a, Quote 10). Negotiating these competing boundaries highlights the complexity of everyday life in self-isolation.

### Maintaining a connection to the outside world

In addition to their sense of duty and a desire to protect others, participants also discussed several ways in which maintaining contact with the outside world could help them preserve the conceptual boundaries they had created and therefore help them adhere to self-isolation. Maintaining contact with the outside world was generally conceptualised in three ways; through social connectivity, tangible practical support and a sense of feeling known to public health authorities.

#### Social connectivity

Maintaining social connectivity was important for mediating the impact of self-isolation on mental health and wellbeing; this was either through socially distanced visits or virtually (Table 3b, Quote 1). Participants also highlighted that maintaining a link with the outside world was particularly important as they were isolating early on in the pandemic. As such, their experience was unique and support from others who were going through the same experience was important (Table 3b, Quote 2). Similarly, survey respondents (37/250, 16%) who agreed or strongly agreed that following self-isolation advice completely would have caused them to lose touch with their friends or family were more likely to report leaving home for higher-contact reasons (35 vs 13%).

#### Practical support

The importance of maintaining a connection with the outside world was also highlighted for practical reasons, such as access to essential supplies including food and medication. At this stage, there were difficulties in accessing online grocery deliveries, as well as financial barriers due to minimum spend for deliveries at some supermarkets. Participants highlighted the importance of having a support network on the “outside” that could help with access to essentials (Table 3b, Quote 3 and 4).

In the survey, grocery delivery was a clear facilitator of adherence. Delivery slots at this time were in short supply and 44 (18%) of 250 self-isolating respondents tried but were unable to secure one. They were distinctly more likely to have left home for higher-contact reasons than the 132 who did get deliveries and the 74 who did not try to (35% vs 10% and 16%, respectively) and – unsurprisingly – specifically for essential shopping (30% compared with 4% and 9%). More generally, 136/250 survey respondents (42%) agreed that they had received help from someone outside their home during their self-isolation period and, compared with others, they were less likely to leave home for higher-contact activities (11 vs 23%).

#### Feeling known

Feeling known to public health agencies also helped participants feel connected to the outside world during their isolation and this supported them to adhere to the guidance. Having the connection with someone in public health agencies, sometimes on a daily basis, helped reassure participants they had not been forgotten. This was particularly relevant for cases, who were sometimes anxious that they might need additional support if their symptoms worsened (Table 3b, Quote 5 and 6).

In some instances, the regular contact from public health agencies resulted in people feeling that they were being monitored. Some participants suggested that the feeling of being known could help people adhere to self-isolation, even though they were not actually being checked (Table 3b, Quote 7).

This was reflected in the survey where, after their first contact with the Health Protection Teams, 77 were contacted on some days and 107 every day during their isolation period (with the remaining 66 reporting no further contact). Of the 184 people who received further contact, 111 (60%) were contacted via text, 119 (65%) by phone and only 25 (14%) via email. Compared with the 73 who were contacted only by phone and/or email, there was some evidence that those whose further contact included text messages were less likely to leave home for higher-contact reasons (11% vs 21%). Text contact was more regular than phone contact, reported as ‘every day’ (rather than ‘some days’) by 79% (50/63) of those who had texts but not calls, 34 % (24/71) of those receiving calls but not texts, and 69% of those who had both texts and calls. However, evidence of association with adherence was stronger for contact by text than for contact every day (supplementary table S4).

### Negotiating competing needs

Participants discussed needs, which acted as barriers to them being able to fully adhere to self-isolation, primarily the practical need to access and store sufficient food. The sudden shift from normal life to the constraints of being a case or contact meant they were unprepared for a two-week isolation period (Table 3b, Quote 8). Some participants’ homes presented additional barriers such that they could not adequately prepare, even if they had known they needed to isolate – for example, not having a big enough fridge to store sufficient food (Table 3b, Quote 9).

### Rational adaptations to mitigate risk

As a way of negotiating these competing needs, participants discussed making rational adaptations to their behaviour. These adaptations were focused on minimising risk of transmission of the virus, while still enabling participants to participate in the behaviours and routines they felt they needed to. For example, if participants did not have access to outside space at home (16% of our survey respondents), they felt they needed to leave isolation so they could exercise outdoors, as this was important for their mental health. However, they purposely did this at specific times of day when they felt confident they would not come into contact with others (Table 3c, Quote 1 and 2).

Similar competing needs were highlighted around pet ownership, specifically dogs. Overall, 43% (108/250) of survey respondents reported having a pet, primarily dogs (28%) and cats (22%). Those who had a pet at home were less likely to report leaving home for higher-contact reasons (10% vs 21%). Welfare concerns over their pets meant that in some cases participants did not fully adhere to self-isolation. However, their decision on *how* to break isolation guidance was based on minimising risk of contact with others, for example walking early in the morning or at places they knew would be quiet (Table 3c, Quote 3 and 4).

#### Over-adherence

In some cases, participants in both the survey and interviews reported over-adhering to the self-isolation guidance, during and after isolation. In the survey a quarter (40/161) of survey respondents who had received advice only to stay inside went beyond that and actually stayed in their room most days – 29 nearly every day, 11 on over half the days – and a further 25 (16%) did so occasionally.

In the interviews, some participants described how their experience of self-isolation had lasting impacts on their perceptions of COVID-19 risk and consequently their behaviour: they felt anxious following their self-isolation and were reluctant to leave the safety of their home (Table 3c, Quote 5 and 6). In some cases, this resulted in over-adherence to COVID-19 guidance. For example, one participant discussed living with their “bubble”, which continued after their isolation period ended (Table 3c, Quote 7).

## Discussion

Understanding how the public make decisions about following self-isolation guidance is important to ensure appropriate provision is in place to support adherence. Our study adds to the growing body of evidence that, despite frequently being reported simply as adherent or not, adherence to self-isolation and the decisions surrounding it are more intricate and often in conflict with activities perceived as essential, such as buying food, exercising outdoors, and dog walking.

The participants in our study understood the reasons for isolating – to reduce the number of contacts they had, and so reduce the risk of transmission. They then used this knowledge to make decisions around how to adhere to the guidance, based on balancing the perceived risk of transmission with maintaining their health and wellbeing (and that of companion animals) during isolation. This was facilitated by a sense of care and connection, balanced with a sense of security provided by the state.

In our study, participants described the impact of their initial contact with public health authorities as resulting in a sudden shift in their identity to become a case or contact. This brought with it an acknowledgement they were now a potential risk to others and embedded within this was a sense of duty to protect those around them. For our participants, this sense of duty to protect others – primarily to save lives – acted as a motivator to adhere to self-isolation. While the majority of survey respondents also agreed that isolation would help protect the NHS, this did not have the same influence over adherence, suggesting participants did not necessarily associate protecting the NHS with saving lives. A sense of duty and desire to protect the community has been identified as a motivator to adhere to self-isolation previously^14^ and is also a key principle in embedding behavioural science into public health campaigns, with emphasis on messages that promote mutual protection and collective solidarity^15^.

The impact of being identified through contact tracing also highlights the importance of the knowledge and expertise of the public health teams doing the contact tracing – specifically being able to offer expert, professional support alongside isolation guidance. Participants in our study found that contact with public health teams helped them feel a connection with authority and gave them a sense of “feeling known”, which resulted in a feeling of security provided by the state; that those in authority cared about their experiences and wellbeing during isolation. This was also reflected in the survey, where regular text contact during isolation was associated with lower risk behaviour. This was particularly important during the early stages of the pandemic, when there was so much uncertainty around the virus and the concept of self-isolation had not yet been embedded in the public consciousness.

Maintaining a connection with the outside world during isolation was also identified as a key motivator to adherence, specifically social connections and practical support. The way in which isolation was enacted by participants was to create a boundary around their living space (home or room within the home) to keep the virus within its confines. However, it is important to be able to maintain a connection with the world outside that boundary, without damaging its integrity. For the participants in our study, this included maintaining social connections, either virtually or socially distanced. Where participants felt they would lose touch with family or friends, they were more likely to leave home during isolation. In addition to social support, practical support was also highlighted by our participants as a key facilitator for adherence to isolation, particularly access to essentials such as food and medicines. This emphasises the importance of providing tangible practical support for people who are isolating, which has been identified previously as a facilitator to adherence^5^.

Using binary measures for adherence has resulted in some studies reporting concerningly low self-reported adherence to self-isolation of 25.0% and 42.5% ^5,16^, whereas other studies have reported much higher levels of 77.8%^17^ and 90.0% ^18^. In our study, strict adherence to isolation (not leaving the home at all) in people directly contacted by a public health team and instructed to isolate was 63%; however when taking into account breaches to isolation that were perceived as lower risk (dog walking, exercise), then adherence was over 80%. Reporting adherence as a binary outcome, without taking into account the complex decisions people are making about their isolation could be problematic and not reflective of reality. Nonetheless, there can be dangers involved in engaging in behaviours perceived as lower risk. In our sample, 10% of people who reported only engaging in lower risk activities still reported being in contact with others within 2 m distance, indoors; a further 10% reported indoor contacts while maintaining at least 2 m distance. Lower risk is not no risk – and if self-isolation is to be used to quickly contain a future infectious disease outbreak, a focus on ensuring people better understand the risk of different activities may be required.

To understand these nuances in adherence, several studies have suggested alternative measures. For example, Fancourt et al^9^ differentiated between ‘complete’ adherence – those who *always* follow *all* the guidance – with ‘majority’ adherence – those who follow *some* of the guidance *some* of the time. In addition, Williams et al^10^ describe the difference between intentionally not following guidance (‘overt rule breaking’) and changing or interpreting guidance to suit individual circumstances (‘subjective rule interpretation’). Denford et al^7^ took this one stage further to explain the complexities of decision-making around adherence to social distancing and self-isolation and identified three patterns of adherence; caution-motivated super-adherence, risk-adapted partial adherence and necessity-driven partial-adherence. For those who partially adhered to guidance, Denford et al found that these decisions were driven by two main factors. For some, decisions were based on personal perceptions of risk: behaviours considered to entail low risk of transmission (i.e. limited or no contact with others) were deemed safe and therefore partial adherence was justified. For others, decisions to break rules were based on tensions between an intention to adhere and a desire to stay safe on the one hand and a need to maintain their mental health and wellbeing or concern over financial responsibilities on the other. In our study, there was little evidence that participants strayed from guidance due to sheer disregard for rules, but rather consciously and thoughtfully to carry out activities that they felt were essential or perceived as low risk such as exercising outdoors or walking their dogs.

### Limitations

Our study is based on a distinct sample of people, who were some of the first COVID-19 cases and their contacts in England, during the first phase of the pandemic response. While this enabled us to gather unique insights into experiences of self-isolation during the early stages of a global pandemic, the sample population is not representative of the wider population. During the early phases of the pandemic response, testing and contact tracing focused on returning holiday makers and travellers and their contacts; consequently, the sample population is primarily White British, of a similar age and from more affluent areas. We attempted to mitigate this by recruiting some interview participants from areas of lower IMD to ensure their experiences were included in the study. However, as the majority of our sample population were from more affluent areas, they may have experienced different barriers to adherence, compared with people for whom access to practical and social support may be more challenging.

The low response rate will have introduced response bias, and the delay between respondents being asked to isolate and inviting them to take part in the study (up to 6 months) will have resulted in recall bias as participants may not have been able to accurately recall their behaviours during the early stages of the pandemic. Recall may have been particularly challenging during the early stages of the pandemic, where there was so much uncertainty and rapidly changing advice and guidance. While this mixed methods study was able to provide some more detailed insights into the behaviours of people asked to self-isolate, our survey questions were not able to fully explore what contacts occurred and why, and it is possible that not all relevant activities and contacts were acknowledged and disclosed. It is also likely that participants who were more adherent were more likely to respond to the survey; concern about disclosing non-adherence or significant breaches of isolation may have discouraged less adherent people from taking part in the study. However, some respondents in our study did disclose instances where isolation guidance was not fully adhered to.

For this analysis, we classified reasons for leaving the home pragmatically as higher or lower contact, as a proxy for potential risk of transmission. However, studies published since we designed the survey have refined our understanding of transmission risk; for example, risk from visiting a supermarket is lower than having people visit your home [19]. Future surveys should focus on the types of activities engaged in and places visited in more detail to relate adherence behaviour to transmission risk more accurately.

## Conclusions

The participants in our study demonstrated they were making rational adaptations to self-isolation guidance, based on a calibration of the risk of transmission, attempting to reduce contact with others as much as possible. Our findings highlighted that these decisions were driven by a sense of duty to protect others. Where isolation was not adhered to, breaches were often for reasons considered essential. The need for adequate practical, financial and social support during isolation has now been well documented; however, our findings highlight the additional impact of contact tracing on identity and feelings of ‘being known’ when asked to self-isolate. This emphasises that isolation cannot be viewed as a single intervention, but should be part of a complete test, trace and isolate process, where all components need to work together to support the desired outcome – reduction in transmission. Better understanding and support for decisions around adherence to isolation during the first phase of pandemic response, when uncertainty and anxiety is unavoidably high, is vital for pandemic preparedness for future emerging infectious diseases to ensure effective containment in the early stages of the response.

## Data Availability

The datasets analysed in this study are available from UK Health Security Agency on reasonable request.

## Acknowledgements

The authors would like to express their gratitude to all the participants who shared their experiences of self-isolation.

G.L., P.C., H.L., M.H., S.D., I.O., C.R., R.R., R.A. and L.Y. are supported by the NIHR Health Protection Research Unit (HPRU) in Behavioural Science and Evaluation at the University of Bristol in partnership with UK Health Security Agency (UK HSA).

L.S., J.R. and R.A. are supported by the NIHR HPRU in Emergency Preparedness and Response at King’s College London in partnership with UK HSA.

L.Y. is an NIHR Senior Investigator and her research programme is partly supported by NIHR Applied Research Collaboration (ARC)-West, NIHR Health Protection Research Unit (HPRU) in Behavioural Science and Evaluation, and the NIHR Southampton Biomedical Research Centre (BRC).

C.R. is affiliated to the National Institute for Health Research Health Protection Research Unit (NIHR HPRU) in Emerging and Zoonotic Infections at the University of Liverpool in partnership with UK HSA in collaboration with the Liverpool School of Tropical Medicine and The University of Oxford, the NIHR HPRU in Gastrointestinal Infections at the University of Liverpool in partnership with UK HSA, in collaboration with the University of Warwick and the NIHR HPRU in Behavioural Science and Evaluation at the University of Bristol, in partnership with UK HSA. C.R. is based at UK HSA.

The views expressed are those of the authors and not necessarily those of the NIHR, the Department of Health and Social Care or UK HSA. The funders had no role in the design of the study, collection, analysis and interpretation of the data, or in writing the manuscript.

## Author contributions

All authors were involved in conceptualising and designing the study. CR, RR and PC analysed the data and CR and RR led the drafting of the manuscript. All authors reviewed the manuscript, approved the final content, and met authorship criteria.

## Patient and public involvement

This was responsive research designed rapidly during the initial response to the COVID-19 pandemic, so it was not possible to involve patients or the public in the development of the study. However, we engaged in preliminary qualitative interviews with a small number of people who had self-isolated during the first few weeks of the pandemic, which were used to inform the development of the survey questions and interview topic guide for the study.

## Patient consent for publication

All participants provided written or oral consent for data to be included in publications.

## Ethics approval

Ethical approval was provided by the NHS Health Research Authority London – Queen Square Research Ethics Committee (20/HRA/2549).

## References

1. Chung S-C, Marlow S, Tobias N, et al. Lessons from countries implementing find, test, trace, isolation and support policies in the rapid response of the COVID-19 pandemic: a systematic review. BMJ Open 2021;11:e047832. doi:10.1136/bmjopen-2020-047832

2. Wilder-Smith A and Freedman DO. Isolation, quarantine, social distancing and community containment: pivotal role for old-style public health measures in the novel coronavirus (2019-nCoV) outbreak. Journal of Travel Medicine 2020; 27(2): 1–4.

3. Buitrago-Garcia D, Egli-Gany D, Counotte MJ, et al. Occurrence and transmission potential of asymptomatic and presymptomatic SARS-CoV-2 infections: A living systematic review and meta-analysis. PLoS Med 2020; 17(9): e1003346. doi. org/10.1371/journal.pmed.1003346

4. Byambasuren O, Cardona M, Bell K, et al. Estimating the extent of asymptomatic COVID-19 and its potential for community transmission: Systematic review and meta-analysis. Journal of the Association of Medical Microbiology and Infectious Disease Canada 2020; 5(4): 223–234.

5. Smith LE, Amlôt R, Lambert H et al. Factors associated with adherence to self-isolation and lockdown measures in the UK: a cross-sectional survey. Public Health 2020; 187: 41-52.

6. Atchison C, Bowman LR, Vrinten C, et al. Early perceptions and behavioural responses during the COVID-19 pandemic: a cross-sectional survey of UK adults. BMJ Open 2021;11:e043577. doi: 10.1136/bmjopen-2020-043577

7. Denford S, Morton KS, Lambert H et al. Understanding patterns of adherence to COVID_19 mitigation measures: a qualitative interview study. Journal of Public Health 2021; doi:10.1093/pubmed/fdab005

8. Thorneloe, R, Clarke, E, Arden, M. Adherence to Behaviours Associated with the Test, Trace, and Isolate System: An Analysis Using the Theoretical Domains Framework. OSF Preprints:10.31219/osf.io/uxbfa

9. Fancourt D, Bu F, Mak H, Steptoe A. A UCL COVID-19 Social Study. Results Release 2020. https://www.covidsocialstudy.org/results (September 2021, xdate last accessed).

10. Williams SN, Armitage CJ, Tampe T, et al. Public perceptions and experiences of social distancing and social isolation during the COVID-19 pandemic: a UK-based focus group study. BMJ Open 2020;10:e039334. doi: 10.1136/bmjopen-2020-039334

11. White R and Van Der Boor C. Impact of the COVID-19 pandemic and initial period of lockdown on the mental health and well-being of adults in the UK. BJPsych Open 2020; 6 (e90): 104 doi: 10.1192/bjo.2020.79

12. Smith LE, Potts H Amlôt R et al. How has the emergence of the Omicron SARS-CoV-2 variant of concern influenced worry, perceived risk, and behaviour in the UK? A series of cross-sectional surveys. OSF Preprints: doi.org/10.31219/osf.io/rpcu2

13. Charmaz, K. (2006) Constructing grounded theory: A practical guide through qualitative research. London: SAGE Publications.

14. Drury, J., Carter, H., Ntontis, E., & Guven, S. T. (2021). Public behaviour in response to the COVID-19 pandemic: understanding the role of group processes. BJPsych open, 7(1).

15. Bonell C, Michie S, Reicher S et al. Harnessing behavioural science in public health campaigns to maintain “social distancing” in response to the COVID-19 pandemic: key principles. Journal of Epidemiology Community Health 2020;74:617–619.

16. Smith LE, Potts HW Amlôt, et al. Adherence to the test, trace, and isolate system in the UK: results from 37 nationally representative surveys. BMJ 2021, 372.

17. Kyle, R.G., Isherwood K.R., Bailey J.W., Davies, A.R. (2021). Self-isolation confidence, adherence and challenges: behavioural insights from contacts of cases of COVID-19 starting and completing self-isolation in Wales. Cardiff: Public Health Wales. Available online: https://phw.nhs.wales/publications/publications1/self-isolation-confidence-adherence-and-challenges-behavioural-insights-from-contacts-of-cases-of-covid-19-starting-and-completing-self-isolation-in-wales/

18. ONS. (2021). Coronavirus and self-isolation after being in contact with a positive case in England: 1 March to 6 March 2021. Available online: https://www.ons.gov.uk/peoplepopulationandcommunity/healthandsocialcare/conditionsanddiseases/bulletins/coronavirusandselfisolationafterbeingincontactwithapositivecaseinengland/1marchto6march2021

19. SAGE. (2021). EMG Transmission Group: Insights on transmission of COVID-19 with a focus on the hospitality, retail and leisure sector, 8 April 2021. Available online: https://www.gov.uk/government/publications/emg-transmission-group-insights-on-transmission-of-covid-19-with-a-focus-on-the-hospitality-retail-and-leisure-sector-8-april-2021

