## Supplementary tables for "Understanding adherence to self-isolation in the first phase of COVID-19 response"

### **Table S1 Exclusions**

| Exclusions based on HPZone data (N) | | |
| --- | --- | --- |
|  | Cases^1^ | Contacts^2^ |
| **Start**: Deduplicated extract of HPZone records | 3580 | 5590 |
| *Exclusions based on HPZone records* |  |  |
| Cases not clearly meeting Covid-19 definition | 18 | 0 |
| Age under 18 | 356 | 928 |
| ^3^Special situations | 3 | 146 |
| ^4^Does not meet survey definition of a contact | 0 | 864 |
| ^5^Not contacted by PHE or contacted too late | 365 | 367 |
| ^6^Event date on or after 10 March 2020 | 45 | 363 |
| ^7^Administrative reasons | 742 | 754 |
| *Exclusions based on external checks* |  |  |
| Newly-discovered duplicates | 3 | 6 |
| No NHS number match | 16 | 34 |
| Not cleared by national opt-out system | 195 | 328 |
| Recorded as died | 15 | 6 |
| **End**: Remaining and invited to participate: N=3616 | 1822 | 1794 |

Notes

^1^ Potential survey cases.

^2^ Potential survey contacts, including contacts of confirmed/possible cases, and ‘exposed persons’ (e.g. returning from a high-risk area).

^3^ Health care workers at supported isolation facilities and specific cruise ship repatriations. These were eligible for other surveys.

^4^ Not a contact by HPZone definition; not monitored by PHE; contact of possible (not confirmed) case; returnee from an affected area but not known to be a contact of a case.

^5^ Too late means at a time when any 14-day follow-up or isolation period would already have been completed.

^6^ Cut-off date set to avoid confusing any advice for a 14-day isolation/monitoring period with national restrictions (“lockdown”) announced on 23 March.

^7^ In pilot sample of 30 (includes four who would have been excluded by preceding criteria, had they been finalised at the time of pilot selection);missing date of birth; missing mobile phone number.

### **Table S2 Characteristics of survey invitees and respondents**

| Characteristic |  | All invitees  N=3616 | Respondents  N=322 |
| --- | --- | --- | --- |
| Case/Contact: N (%) | Confirmed case  Possible case  Contact | 350 (9.7%)  1472 (40.7%)  1794 (49.6%) | 52 (16.2%)  91 (28.3%)  179 (55.6%) |
| Age: median (IQR) | Years | 42 (29–54) | 48 (35–58) |
| Gender | Female  Male  Missing/prefer not to say | 1862 (51.5%)  1667 (46.1%)  87 (2.4%) | 200 (62.1%)  118 (36.7%)  4 (1.2%) |
| Index of multiple deprivation (IMD) quintile of local area:^1^ N (%) | 1 (most deprived)  2  3  4  5 (least deprived)  Missing | 410 (11.3%)  631 (17.5%)  744 (20.6%)  785 (21.7%)  937 (25.9%)  109 (3.0%) | 24 (7.5%)  44 (13.7%)  80 (24.8%)  75 (23.3%)  89 (27.6%)  10 (3.1%) |
| Ethnic group: N (%) | White^2^  All other ethnic groups^3^  Missing/prefer not to say | Not available  Not available  . | 287 (89.1%)  33 (10.2%)  2 (0.6%) |

^1^ IMD defined for the local area (lower layer super output area) of participant’s postcode

^2^ White (British/Irish/Other)

^3^ Asian, Black/Black British, Chinese, Mixed, Other

### **Table S3 Interviewee characteristics**

| Participant | Group | Age group | Gender | IMD decile |
| --- | --- | --- | --- | --- |
| 1 | Contact | 45 to 69 | Female | 5 |
| 2 | Case | 45 to 69 | Female | 6 |
| 3 | Case | 45 to 69 | Male | 5 |
| 4 | Case | 45 to 69 | Female | 6 |
| 5 | Case | 45 to 69 | Female | 6 |
| 6 | Contact | 45 to 69 | Male | 6 |
| 7 | Contact | 45 to 69 | Female | 5 |
| 8 | Case | 45 to 69 | Female | 6 |
| 9 | Contact | 25 to 44 | Female | 5 |
| 10 | Contact | 25 to 44 | Female | 4 |
| 11 | Case | 45 to 69 | Female | 5 |
| 12 | Case | 45 to 69 | Female | 1 |
| 13 | Case | 25 to 44 | Female | 2 |
| 14 | Contact | 25 to 44 | Female | 2 |
| 15 | Contact | 45 to 69 | Female | 7 |
| 16 | Contact | 45 to 69 | Female | Not available |

### **Table S4 62 potential predictors of adherence to self-isolation advice**

|  |  | **Going-out behaviour: N of respondents^2^** | | | | | |  |
| --- | --- | --- | --- | --- | --- | --- | --- | --- |
|  |  | **Predictor = “No”** | | | **Predictor = “Yes”** | | |  |
| **Q** | **Meaning of ‘Yes’ in binary predictor^1^** | **None^3^** | **Low^4^** | **High^5^** | **None^3^** | **Low^4^** | **High^5^** | **P^6^** |
| n/a | Survey phase 2 | 51 | 23 | 20 | 107 | 28 | 21 | 0.073 |
| n/a | Case (confirmed/possible) | 92 | 31 | 21 | 66 | 20 | 20 | 0.630 |
| n/a | More deprived local area (IMD quintiles 1-3) | 74 | 30 | 17 | 78 | 20 | 22 | 0.250 |
| 5 | Very/extremely worried about coronavirus (at time of survey) | 112 | 38 | 30 | 46 | 13 | 11 | 0.898 |
| 7 | Advised to stay in room | 123 | 47 | 34 | 35 | 4 | 7 | 0.060 |
| 8 | PHE follow-up messages every day | 89 | 26 | 28 | 69 | 25 | 13 | 0.233 |
| 9a | Follow-up messages by text | 47 | 11 | 15 | 70 | 29 | 12 | 0.070 |
| 9b | Follow-up messages by email | 103 | 33 | 23 | 14 | 7 | 4 | 0.605 |
| 9c | Follow-up messages by phone | 40 | 16 | 9 | 77 | 24 | 18 | 0.771 |
| 10 | Follow-up messages were very/extremely useful | 74 | 24 | 15 | 43 | 16 | 12 | 0.720 |
| 11 | PHE gave all the information I needed | 57 | 20 | 14 | 101 | 31 | 27 | 0.861 |
| 12/13 | Had fever, dry cough or breathing difficulty | 91 | 38 | 26 | 67 | 13 | 15 | 0.092 |
| 15 | I think I probably/definitely had coronavirus | 89 | 23 | 20 | 69 | 28 | 21 | 0.333 |
| 16 | I had a positive test for coronavirus | 120 | 34 | 28 | 38 | 17 | 13 | 0.320 |
| 17 | Lived alone | 131 | 45 | 31 | 27 | 6 | 10 | 0.295 |
| 18a | Shared house: children aged 0-4 | 113 | 43 | 28 | 18 | 2 | 3 | 0.223 |
| 18b | Shared house with children aged 5-17 | 85 | 32 | 24 | 46 | 13 | 7 | 0.368 |
| 18c | Shared house with adults aged 18-69 | 7 | 4 | 1 | 124 | 41 | 30 | 0.677 |
| 18d | Shared house with adults aged >=70 | 122 | 39 | 30 | 9 | 6 | 1 | 0.259 |
| 19 | Shared house with family | 15 | 4 | 2 | 116 | 41 | 29 | 0.798 |
| 20 | Home had shared bathroom facilities | 17 | 8 | 6 | 114 | 37 | 25 | 0.534 |
| 21 | Home had a room to live and sleep in | 36 | 5 | 8 | 95 | 40 | 23 | 0.070 |
| 22 | Home had access to any outside space | 29 | 1 | 9 | 129 | 50 | 32 | 0.003 |
| 23 | Had any pets that lived in home | 86 | 26 | 30 | 72 | 25 | 11 | 0.059 |
| 25 | Others in household self-isolated with me | 45 | 10 | 10 | 85 | 35 | 20 | 0.296 |
| 30 | I tried but was unable to arrange grocery delivery | 136 | 45 | 25 | 22 | 6 | 16 | 0.001 |
| 32a | Difficulty: no room I could use to stay in on my own | 117 | 43 | 25 | 14 | 2 | 6 | 0.125 |
| 32b | Difficulty: I had to look after other people | 112 | 36 | 27 | 19 | 9 | 4 | 0.644 |
| 32c | Difficulty: family wanted/needed to talk to or see me | 114 | 37 | 23 | 17 | 8 | 8 | 0.178 |
| 32d | Difficulty: very ill; family members had to look after me | 124 | 43 | 24 | 7 | 2 | 7 | 0.009 |
| 36 | High-risk household member | 105 | 37 | 25 | 23 | 8 | 6 | 0.966 |
| 38 | At time of PHE contact, worked or studied outside home. | 62 | 22 | 17 | 96 | 29 | 24 | 0.867 |
| 44a | Following advice: would have lost touch with friends/family | 141 | 44 | 28 | 17 | 7 | 13 | 0.007 |
| 44b | Not following advice: friends/family would have disapproved | 35 | 13 | 13 | 123 | 38 | 28 | 0.420 |
| 44c | Not following advice: could have been in trouble with the police | 87 | 32 | 22 | 71 | 19 | 19 | 0.611 |
| 44d | Following advice: “would have helped save lives” | 17 | 10 | 11 | 141 | 41 | 30 | 0.025 |
| 44e | Following advice: “would have helped protect the NHS” | 20 | 8 | 6 | 138 | 43 | 35 | 0.832 |
| 44f | If I caught coronavirus: “I may have become very ill” | 55 | 15 | 14 | 103 | 36 | 27 | 0.777 |
| 44g | If I caught coronavirus: severe impact on my family’s wellbeing | 60 | 20 | 19 | 98 | 31 | 22 | 0.613 |
| 44h | Following advice: more conflict with people I was living with | 136 | 48 | 34 | 22 | 3 | 7 | 0.194 |
| 44i | Leaving home: I could have passed coronavirus to someone | 20 | 2 | 8 | 138 | 49 | 33 | 0.051 |
| 44j | Leaving home: I could catch coronavirus | 43 | 18 | 9 | 115 | 33 | 32 | 0.348 |
| 44k | Following advice: negative impact on how much money I had | 128 | 43 | 32 | 30 | 8 | 9 | 0.712 |
| 44l | Following advice: would have missed important religious activities | 147 | 50 | 37 | 11 | 1 | 4 | 0.287 |
| 44m | Received help from someone outside my household | 66 | 22 | 26 | 92 | 29 | 15 | 0.042 |
| 44n | Self-isolation made my physical health worse | 108 | 34 | 18 | 50 | 17 | 23 | 0.015 |
| 44o | Self-isolation made my mental health worse | 81 | 30 | 11 | 77 | 21 | 30 | 0.005 |
| 44p | Self-isolation made my physical health better | 142 | 45 | 35 | 16 | 6 | 6 | 0.623 |
| 44q | Self-isolation made my mental health better | 150 | 49 | 39 | 8 | 2 | 2 | 1.000 |
| 44r | I enjoyed spending more time at home during self-isolation | 103 | 32 | 24 | 55 | 19 | 17 | 0.715 |
| 45 | Belief: >=65% of others fully followed PHE's advice (at that time) | 65 | 17 | 19 | 93 | 34 | 22 | 0.437 |
| 46 | Belief: >=65% of others fully follow Govt guidance (at time of survey) | 104 | 31 | 25 | 54 | 20 | 16 | 0.727 |
| 48c | Someone else helped care for pet(s) | 55 | 24 | 8 | 17 | 1 | 3 | 0.046 |
| 50 | Possible depression (PHQ-9 score >=10) "over last 2 weeks" | 131 | 43 | 30 | 27 | 8 | 11 | 0.324 |
| 51 | Possible PTSD (PC-PTSD5 score >=4) "over past month" | 147 | 48 | 37 | 11 | 3 | 4 | 0.773 |
| 52 | Possible anxiety (GAD-7 score >=10) "over last 2 weeks" | 137 | 47 | 32 | 21 | 4 | 9 | 0.150 |
| 53 | Moderately/very angry about self-isolation "over last 2 weeks" | 152 | 46 | 35 | 6 | 5 | 6 | 0.022 |
| 54 | Age 50 or over | 97 | 22 | 27 | 61 | 29 | 14 | 0.041 |
| 55 | Male | 108 | 34 | 22 | 49 | 17 | 19 | 0.208 |
| 56 | Ethnic group: non-White (Asian, Black/Black British, Chinese, Mixed, Other) | 136 | 50 | 37 | 21 | 1 | 3 | 0.044 |
| 57 | Education: degree level or above | 49 | 16 | 8 | 108 | 35 | 32 | 0.363 |
| 59 | Any long-term illness/health problem/disability (at that time) | 122 | 40 | 31 | 36 | 11 | 10 | 0.952 |

^1^ See copy of survey in supplementary information for full question wordings and response options. For Qs 11, 44a–44r, Yes = agree/strongly agree with statement.

^2^ Of 250 respondents who had been advised to self-isolate (stay at home or stay in room). N in analysis is less than 250 if the potential predictor was only applicable to a subset: Qs 9–10 (if any follow-up messages were received); Qs 18-21, 25, 32a–32d, 36 (if living with one or more other people); Q 48c (if had a pet at home); or if data was missing (Qs 25, 36, 55–57 and deprivation only).

^3^ Did not leave home for any reason.

^4^ Left home for lower-contact reasons (exercise and dog-walking) ONLY.

^5^ Left home for any higher-contact reason (possibly in addition to lower-contact reasons).

^6^ Fisher’s exact test

The following survey questions were not considered as potential predictors in this analysis: Q6, 24, 58 (overlap with other factors considered); Q14, 26–29, 33–35, 47, 48a–b (primary or other adherence behaviours); Q39–41 (too few responses in smaller category); Q37, 42–43, 49 (views about future behaviour).
